# A Decolonial Exploration of Stakeholder Perspectives on Cameroon’s Expanded Programme on Immunisation (EPI): A Critical Qualitative Inquiry

**DOI:** 10.64898/2026.08.18.26360686

**Authors:** Oben Pamela Besong, Calvin Tonga, Martha Ndiko Ngoe, Luchuo Engelbert Bain, Nadia Fazal

**Affiliations:** Expanded Programme on Immunisation, Delegation of Public Health, Southwest Region, Cameroon; Liverpool School of Tropical Medicine, U.K; Oxford University, U.K

## Abstract

Despite significant progress in reducing vaccine-preventable diseases, Cameroon’s Expanded Programme on Immunisation operates within structures shaped by colonial history. Overreliance on external donor funding, centralised governance, and limited recognition of local knowledge raise concerns about equity, local ownership, and programme sustainability, particularly as the country plans for donor transition and self-financing. This study, aligned with the decolonising global health movement, examines how colonial legacies shape stakeholders’ experiences within the EPI and proposes practical steps towards a more locally owned immunisation programme. A qualitative case study was conducted in the Southwest Region of Cameroon from June to July 2025, comprising fifteen online semi-structured interviews in English with selected stakeholders (regional and district EPI managers, civil society members, and community leaders). Interviews were audio-recorded, transcribed verbatim, and analysed thematically using Clarke and Braun’s six-step framework in NVivo version 11. Participants identified subtle colonial influences, including centralised decision-making, donor-driven priorities, pay disparities favouring international actors over local staff, and the marginalisation of local and traditional knowledge. The COVID-19 response was frequently cited as an example of inequity, with Western biomedical approaches prioritised over locally led solutions. Major structural issues included heavy reliance on external funding, outdated colonial-era training curricula, centralised governance, and a lack of local vaccine manufacturing capacity. Despite these issues, participants recognised the significant technical and financial support from international organisations. They proposed concrete pathways for decolonisation, including decentralised governance, participatory programme design, regulation and integration of traditional medicine, community engagement, domestic resource mobilisation, leveraging Cameroon’s emerging universal health coverage to reduce donor dependence, and investing in local vaccine production. Conclusion: Colonial legacies continue to influence Cameroon’s EPI, undermining local ownership and self-determination, even when external support is effective. Achieving decolonisation requires multifaceted efforts to strengthen domestic financing and governance, empower local stakeholders, and legitimise local knowledge alongside biomedical approaches. Policymakers should embed local ownership, governance reforms, and local capacity building in transition strategies while donor funding persists, ensuring immunisation gains are sustained beyond external support. These insights provide a context-specific roadmap for developing a sustainable, equitable, and locally driven immunisation programme in Cameroon and other countries facing similar donor transitions.

## Introduction

The EPI is a global health initiative launched by the World Health Organisation (WHO) in 1974 to combat VPDs [1]. Over the past 50 years, the EPI has significantly reduced VPDs among children in Sub-Saharan Africa by strengthening national immunisation programmes and has played a key role in eradicating diseases such as smallpox. Despite notable progress, achieving global vaccination targets remains challenging, shaped by technical, social, political, and historical factors [2]. This study examines these challenges in Cameroon’s EPI through a decolonial lens, considering the historical origins of immunisation, governance, programme financing, and trust.

Colonisation results from power imbalances where one group dominates and subjugates others to exploit and extract wealth. During the colonial era, immunisation was not only used as a preventive health measure but also as a means of exerting control over colonised states [3]. In Central Africa, infections like smallpox and sleeping sickness were controlled through widespread vaccination of indigenous populations, while exploiting the local resources of colonies and advancing scientific research, ultimately resulting in both benefits and detriments for colonised nations [4]. Furthermore, practices such as coercive vaccination policies, lack of informed consent, and the systemic neglect of the safety and well-being of indigenous populations during clinical trials have contributed to mistrust in healthcare systems. This historical mistrust complicates current vaccination initiatives and underscores the enduring impact of colonial practices on public health perceptions to date [4]. In contemporary discourse, colonial tendencies may appear in subtle and hidden forms, framed as charity or aid [3]. These can occur at various scales and levels, even indirectly through unjust structures and systems. Scholars have pointed to the COVID-19 pandemic as a case in point where high-income countries secured a disproportionate share of the earliest global vaccine supply, while the COVAX mechanism intended to guarantee more equitable access fell short for lower-income countries [5–7]. By December 2021, high-income countries had vaccinated roughly 70% of their populations, compared with about 4% in low-income countries [7]. Scholars have characterised this disparity as a pivotal illustration of how colonial-style hierarchies persist within global health governance. This systemic exclusion, compounded by the exploitation of local infrastructure for external benefit, reinforces historical patterns wherein international assistance prioritises donor interests over the sovereign health needs of the Global South [8].

Cameroon in Central Africa has a multifaceted colonial history dating back to French and British colonisation. This dual influence has significantly shaped the country’s social, cultural, political, and health dynamics. As a result, Cameroon grapples with challenges and disparities rooted in this historical context that affect the country’s development and social cohesion [9]. Nationally, routine immunisation coverage has reportedly remained below EPI targets: a 2018 Demographic and Health Survey found that only 42% of Cameroonian children were fully immunised [10], and as of 2023 only 54% of Cameroonian health districts achieved at least 80% coverage for the third dose of the pentavalent vaccine, below the coverage levels reported in several neighbouring Central African countries [11]. Several authors have associated low immunisation coverage in Cameroon with population vaccine hesitancy [12,13]. As defined by WHO, vaccine hesitancy refers to the reluctance or refusal to vaccinate despite the availability of vaccines [14]. In Cameroon, vaccine hesitancy is perceived to have originated from awareness of the coerced vaccination practices implemented by the French during the colonial era [4]. The hesitation of the Cameroonian population to receive COVID-19 vaccines and their preference for traditional remedies during the pandemic, despite warnings from the government and WHO about the lack of scientific evidence, highlight a greater appreciation for traditional remedies over modern medicine [15,16]. However, the undermining of traditional remedies by the government and international organisations draws parallels to colonial practice, where modern medicine was forced to replace traditional remedies [9,13–17]. Cameroon, the EPI is primarily funded by international organisations such as the Global Alliance for Vaccines and Immunisation (GAVI) [18]. These organisations also play a crucial role in shaping policies and strategies for the programme, such as the rollout of new vaccines [19]. However, while international financial support has led to significant achievements, including Cameroon attaining polio-free status in 2020 [20], funding challenges exist. According to Amani et al. [19], heavy reliance on external funding is characterised by insufficient alignment with local needs, low rates of local fund and resource mobilisation, undermining local ownership in immunisation.

Diverse stakeholders (administrative, community, health, religious, and multi-sectoral actors) play an important role in the acceptability, implementation and sustainability of immunisation programmes [21], [22] For instance, Community Health Workers (CHWs) play a central role in community mobilisation for vaccination and the surveillance of VPDs [23]. On the other hand, insufficient engagement of vital stakeholders in decision-making processes can lead to disengagement and apathy towards immunisation initiatives, thus limiting programme effectiveness and performance [24]. Research shows that local stakeholders, like Civil Society Organisations (CSOs), are often underrepresented in development initiatives that concern their communities, reflecting a continuation of the centralised governance system inherited from colonial administration [25], [9] This underrepresentation in health decision-making is particularly pronounced in vertical programmes with top-down approaches, undermining the reflection of the communities’ needs and preferences in health interventions.

Scholars argue that recognising and addressing remnants of colonisation, such as power structures that favour dependence on aid and the underrepresentation of local communities in decision-making, is essential for addressing public health challenges and fostering local ownership [26], [4] Recently, decolonisation discourse has gained momentum in Global Health (GH) practice as an approach that seeks “to remove all forms of supremacy within all spaces of global health practice, within countries, between countries, and at the global level”. Given the historical context, lingering distrust of vaccines, financial dependence on international organisations, and top-down governance structures in Cameroon, there is a need for a decolonial lens to identify and address GH inequities, repair historical trust, and shift decision-making power to local communities [27]. To our knowledge, there is a gap in the literature on decolonial studies of immunisation programmes in Cameroon. This study will therefore provide insights into stakeholders’ (regional and district EPI managers, civil society actors, and community influencers) knowledge and perceptions of colonisation and decolonisation within the EPI in Cameroon, uncovering findings that will inform EPI policies that promote equity, local ownership, and sustainability.

## Methods

### Study Design

A qualitative design using thematic analysis was employed to explore stakeholders’ perceptions of colonisation and decolonisation. Specifically, the inquiry examined stakeholders’ knowledge of colonisation, experiences and challenges, power dynamics, and partnerships and opportunities for decolonising the immunisation programme. A two-minute read of a fictional vignette was included at the start of the questionnaire to illustrate colonisation and decolonisation and to elicit participants’ memories and perceptions [28]. Data were collected through semi-structured interviews conducted via Microsoft Teams.

### Overview of the EPI

The EPI is a key health programme in Cameroon that offers free vaccination services to prevent, control, and eliminate VPDs. The programme started as a pilot project carried out by the Organisation for the Coordination and Cooperation of the Fight against Endemic Diseases in Central Africa (OCEAC) in 1976 and has evolved to include the delivery of vaccines to children, adolescents, women of child-bearing age, and adults aged 18 years and above [29]. The government and developmental partners like GAVI provide funding, vaccines, and other resources for the programme’s implementation [18]. The Southwest EPI Regional Coordination is an arm of EPI for decision-making and implementation. It is headed by a regional coordinator who oversees the management and coordination of the regional EPI team and coordinates policy implementation across the region’s 21 health districts. Population size, diversity, and health priorities are important factors in allocating funding for EPI interventions. The regional EPI coordinator reports directly to the Regional Delegate of Public Health, who coordinates all regional health programmes, and the EPI Permanent Secretary at the national level [29].

#### 3.2.1 Study period

We conducted 15 online Key Informant Interviews (KIIs) from June 8 to July 28, 2025.

#### 3.2.2 Study Population

Study participants included:

1. **The Regional Delegate (n=1):** Oversees all regional health programmes.
2. **Regional Unit Heads (n=3):** Responsible for surveillance, cold chain, SBCC, and finance.
3. **Chiefs of Health Districts (n=7):** Managers overseeing the 21 health districts.
4. **Civil Society Actors (n=2):** Representatives from local NGOs.
5. **Opinion Leaders (n=2):** Key figures providing cultural and community insights.

### Participant Selection Sampling

Purposive sampling was used to select fifteen (15) participants, ensuring gender representation and accounting for years of experience (>=3) to provide diverse perspectives and rich insights. A sample of fifteen (15) was considered convenient for achieving data saturation, as reported in the literature [30].

### Data Collection

Study participants were contacted at least 10 days before their interviews by email, phone, or in person. This approach enabled us to reach participants more efficiently. During these communications, each participant received an information letter (Appendix 2) explaining the study’s purpose and their rights. If an email went unanswered within 5 days, a follow-up call was made to confirm contact or to schedule an in-person discussion for clarification. Thereafter, participants had 5 days to review materials and ask questions before signing and submitting their consent forms (Appendix 3) to the principal investigator. Overall, 20 potential participants were approached, yielding a response rate of 75%.

Fifteen online interviews were conducted in English, each lasting 45–90 minutes. During the online interview preparatory phase, participants’ availability, internet access, and ability to use Microsoft Teams were verified through WhatsApp discussions and phone calls. A semi-structured, one-on-one interview format (see Appendix 1) was preferred as it facilitates in-depth discussions of sensitive topics such as colonisation, which may be difficult in a group setting. This approach also helped maintain participants’ focus during the interviews and limit unnecessary divergence [31,32]. The interview guide comprised open-ended questions on participants’ experiences and perceptions regarding (1) experiences of colonisation within the EPI in Cameroon; (2) the challenges associated with efforts to decolonise the EPI in Cameroon; (3) opportunities for advancing decolonisation within the EPI in Cameroon. All interview data were stored on a password-protected computer for security purposes.

Finally, the principal investigator used a reflexive journal to record feelings and assumptions before, during, and after the study [33]. It was structured as (1) a log of evolving perceptions, (2) a log of day-to-day procedures, (3) a log of methodological decisions, and (4) a log of day-to-day introspections and how these influenced the findings.

### Data Analysis

Transcripts were automatically generated in Microsoft Teams following interviews. After reviewing the transcripts and comparing them with the recordings, minimal grammatical corrections were made to improve readability and understanding. The proofreading process was also used to become familiar with the collected data [34]. NVivo version 11 software and Clarke & Braun’s six-step methodology (1) familiarising with the data, (2) generating initial codes, (3) collating codes into themes, (4) reviewing themes, (5) defining and naming themes, and (6) producing a report were used to analyse the data [34]. After three initial interviews, transcripts were coded to identify preliminary themes, which the research team discussed, leading to a refinement of the interview guide used with the other participants.

### Data Validity

To ensure trustworthiness and data validity, participant recordings were transcribed verbatim with very few grammatical corrections to improve comprehension. A robust data trail, including recordings and transcripts, was kept, alongside a logbook detailing the principal investigator’s thoughts and perceptions throughout the study.

### Ethical Considerations

Ethical approval was obtained from the Southwest Regional Ethics Committee for Human Health Research in Cameroon (Appendix 4) and the LSTM Master’s Review Panel (Appendix 5). Participants received information forms by email or in print, outlining the study’s purpose, their right to withdraw, and confidentiality measures to protect their identities. We obtained consent from all fifteen (15) participants, who signed consent forms confirming their participation in the online interviews. All transcripts were carefully reviewed to remove any identifiers, including participants’ names, locations, and educational levels. To mitigate emotional distress during the interviews, we informed participants of the potential for distress, and only those who agreed to take part were included. During the interviews, we also showed empathy through nods and local cues to demonstrate that participants’ emotional expressions (mostly raised voices, anger, and worry) were valid.

### Positionality of the Principal Investigator

The Southwest is one of two English-speaking regions in predominantly francophone Cameroon and has historical ties to Britain. Since 2016, the region has been embroiled in a socio-political crisis that began as protests against marginalisation and later escalated into conflict between state and non-state actors. The ongoing crisis and my familiarity with the research participants created a comfortable atmosphere for discussing a sensitive topic such as colonisation. Participants expressed hope for policy changes that would support more equitable and locally owned programmes, while taking their priorities into consideration.

I am a Cameroonian woman from Cameroon’s English-speaking Southwest region and hold a Doctorate in Pharmacy from the University of Douala. The decolonial exploration of stakeholder perspectives on the Expanded Programme on Immunization in Cameroon emerged during my master’s degree in Global Health at the Liverpool School of Tropical Medicine, UK, in 2025. At that time, I had recently learned about power in global health and began reflecting on how my role as an immunisation manager in Cameroon had made me, unintentionally, a perpetrator, a victim, or both, of colonial practices within the health system. To better understand my role and those of other stakeholders in the Southwest Regional EPI, I undertook this study and drew on insider– outsider dynamics to reflect on my positionality, thoughts, emotions, and how they interacted with the research. Recognising that stakeholders, including me, might not have had a thorough understanding of these topics [35], [36], I included a two-minute fictional vignette at the beginning of the interview guide to illustrate colonisation and elicit stakeholders’ perspectives [28].

## Results

The study findings are presented under three main themes: perceived colonial experiences in the EPI in Cameroon; challenges associated with efforts to decolonise the EPI in Cameroon; and opportunities for advancing decolonisation within the EPI in Cameroon.

### Perceived colonial experiences in the EPI

#### Knowledge of colonisation

Most participants acknowledged the lingering colonial tendencies in Cameroon and the EPI, expressed mainly through a lack of freedom in decision-making over foreign interests. Participants recognised that their experiences of colonisation were backed by knowledge of Cameroon’s colonial history.

**P1:** “But gradually you can see, and then you take stock of what is happening. You will find a colonial tendency. Do not forget that Cameroon is one of the countries that were colonised, as some of us studied. We had stories and have seen examples of this colonialism that transcends time.”

However, unlike colonial periods, when practices were outright brutal, recent practices were described as “subtle or indirect.”

**P15:** “But I also think colonialism is not like before.”…I think that that’s the worst aspect of colonisation, where you are educated. You’re educated to know what to do, but someone is telling you what to do. It’s worse than it. Initially, those people were not educated. They did not know what to do. Now we are educated. We know what to do, but we do not do it because we have to take from somewhere else. Even though it is not working, we fear losing their money. I think that colonial tendency from the beginning is still there, but we have just made it less. It is becoming subtle and more civilised. Yes. It has been improved so that even educated people can be captured in the colonial tendencies.”

### Positive Expressions of Dependence on Global North Funding and Technical Support

Contrarily, a few stakeholders expressed colonisation positively, indicating that financial and technical influences from colonial forces are necessary and have contributed to reducing morbidity and mortality from vaccine-preventable diseases.

**P6: “**I believe that colonial influences were present in Cameroon…during the pandemic. Without the influence of foreign bodies, we could have recorded more complicated cases.” However, this reliance on external support often masks deeper systemic issues, where global health practices continue to prioritise Western paradigms over local expertise and indigenous knowledge [37].

#### Preferential Treatment of Expatriates Over Local Actors

GN actors expressed having experienced preferential treatment over local actors, such as better pay for the same job opportunities and prioritising GN actors for specific funding opportunities.

**P2:** “So we did the same work, the same number of days we worked together, but what I received was not even 1/4 of what he had. And he was treated more favourably, yeah.”

**P8:** “Even if in situations where they have to implicate. The private sector and civil societies are involved in immunisation; most civil societies that get these contracts are global civil societies. Based in the developed world, then? They are now working with their subcontractors. Those on the ground.”

#### Experiences during the COVID-19 Pandemic

The most frequently mentioned colonial experiences were during the COVID-19 pandemic, when stakeholders felt that, amidst uncertainty, Western approaches, such as vaccination, were portrayed as superior and prioritised on local practices.

**P9:** “*Yes, I saw it during the COVID-19 pandemic. We had a disease that we had never heard of, never seen.* It *hit us hard with little time to even understand what the disease is all about. This is the time to situate it in our context. In Africa, and Cameroon, we had numerous protocols that we had to follow. We had measures that we had to take. We were not very sure if these worked in our context…and we began to roll out without giving time to understand what works in our context, strategise, and choose the right vaccine contextual to our population.”*

**P5:** “During that period, we had a handful of people from Africa who came up with remedies, but after a while, we noticed that these remedies died out because. They were not being portrayed, and some people were saying no. If we portray them, Europe is going to have a crackdown on us; they’re not going to do this or do that. So it is like the impression that I personally have is that they were being intimidated to be silent because it was seen as a market coming from Europe. That was how we saw it. So why is it that the people in Africa? Those who came out with remedies concerning COVID-19 were not being highlighted on the international platforms because when there is a global crisis, it is a collective issue. Solutions should not come only from Europe. Solutions can also come from Africa…we failed to look at our local herbs.”

Controversies surrounding the COVID-19 pandemic contributed to vaccine hesitancy by generating doubt about the objectives of the vaccines among the African population.

**P5:** “There was much misinformation in the COVID factor. You realise that most people were fleeing because most of this information came from Europe. The misinformation came from Europe, but Europe was sending the vaccines to Africa…So most people do not take it.”

### Challenges associated with the EPI

#### Predominance of external funding and influence on decision-making

According to stakeholders, GN organisations funding the EPI influence the programme’s policies and decisions without considering local contexts.

**P9:** “There are a lot of activities related to EPI, even our logistics, transportation, and everything, and very little of these activities are sponsored by the state in the end. Whatever the sponsors want in terms of the implementation methods and delivery of the results has to align with their own demands.”

**P1:** *“About 95% of all our activities have been sponsored by partners, and like I said, these partners defined the processes…Sometimes I feel that the central level has its hands tied to an extent, which means that they are also a victim of the colonialism which is taking place…”*

#### Lack of local vaccine production

Stakeholders perceive the lack of local production and quality control of vaccines used in the EPI as favouring dependence on GN funding.

**P15:** *“And we are in this tropical zone where this disease is affecting us here in the sub-Saharan Africa…You cannot produce a vaccine here, and you test it out of the country in a country where maybe those people do not even have malaria.”*

#### Curriculum and training

Stakeholders report that GN influences the use of outdated curricula in education.

**P11:** “We also have this part of the colonial curriculum for our many schools…as I earlier said, health workers don’t know because they were not trained, so it is part of that curriculum. Nothing has been updated in that curriculum. And so it’s time to see how we revisit the existing curriculum to update it to today’s world.”

#### Centralised governance structures

The centralised governance structures used by the EPI were seen as colonial, limiting local buy-in and ownership.

**P12:** “…that normally, centralising everything…I’m talking about the French system, where everything is centralised. The colonial mentality that they are up there and you are down there and they decide on everything…So, centralisation may be due to what we inherited from the colonial masters.”

### Opportunities for advancing decolonisation within the EPI

Stakeholders identified specific approaches to promoting local ownership and valuing community opinions.

#### 4.3.2 Decentralisation

Stakeholders emphasised the importance of implementing decentralised governance to promote local ownership.

**P7:** “I’m talking about our system today. We can have decentralisation, federalisation, or any system that gives proper representation to communities. We will go a long way in reducing some of these inaccessibilities because once a community has proper representation, I think it would be easy for their needs and their pleas to be taken into consideration. So, a complete overhaul of the status quo of the current administrative setup.”

##### Regulation of Traditional Practices

Regulation of the traditional sector and collaboration between the EPI and traditional practitioners were expressed as practices that will favour local ownership and reduce vaccine hesitancy.

**P9:** “We are going to regulate that particular sector to see how we can add traditional medicine…It will improve the referral system between traditional medicine and modern medicine. Yes, eventually, when the regulatory aspect has been done, then the partnership with the traditional authorities will also be smoother.”

##### Community Participation and Ownership

Greater involvement of community representatives, civil societies, and community health workers in EPI interventions was seen as an effective way to ensure local representation in EPI decision-making.

**P15:** *“We realise that the Community is like the backbone now in EPI activities because we communities are involved, have community leaders, and the CHWs who already understand them*.

*Community, and they know where to go to make sure that activities are being carried out and that they are being carried out successfully”*

##### Local resource mobilisation

Mobilising resources from various sectors, such as councils, philanthropists, and health facilities, will reduce reliance on external funding.

**P9:** “…at the level of the implementation facilities for them to be able to tailor and invest the revenue generated within their facilities back into EPI implementations. This revenue also belongs to the state, so it is also able to leverage this revenue to continue implementing EPI activities.”

**P6:** “*Now, in terms of funding, the local authorities, like the mayors of different kinds, should also see the need for them to include vaccination in their budgets.”*

##### Local vaccine production

Local vaccine production and quality control were highlighted as necessary to reduce dependence on the GN and provide better quality vaccines to the population.

**P8:** “One big opportunity I see is the African Vaccine Manufacturing Accelerator. And how can we maximise this opportunity? It is for African leaders to get involved…That’s one of the ways. If we begin to take ownership.”

**P15: “**We are better placed to produce these vaccines here and test them here in the country before usage…So I think that by the time we open our own pharmaceutical company here, where we can produce and test them here, we’ll be able to come up with better vaccines. That will have less. After effect, they usually call AEFI.”

##### Universal Health Coverage (UHC)

UHC was seen as a tool for local mobilisation of funding for immunisation.

**P13:** “It’s true that it has been made completely free, that vaccination is free in Cameroon, but I’m sure that if universal health coverage is put in place in a way that all the Cameroonians understand, they should put in something. To take care of their children, and if that is well explained to them, we can generate all the resources locally…”

## Discussion

Our findings provide insights relevant to perceived experiences of colonisation, challenges and opportunities in relation to the EPI Cameroon.

### Perceived experiences of colonisation in the EPI Cameroon

#### Knowledge of colonisation

Our findings suggest that colonial practices continue to operate within the EPI, decades after former colonies gained administrative independence. This ongoing influence aligns with Mbembe’s 2017 texts, which discuss how modern forms of exploitation and dominance persist within global systems [38]. Moreover, stakeholders indicated awareness of the historical foundations of colonisation in Cameroon, largely due to primary and secondary school study of Cameroonian history. This educational background resulted in a strong knowledge base of colonial practices and an understanding of the implications of colonial practice for health programmes and the local population. Here, the connection to key educational theories becomes evident, particularly Paulo Freire’s concept of critical consciousness. Freire emphasises the necessity for individuals to understand their sociopolitical environment in order to confront oppressive systems [39]. In addition to formal awareness, most stakeholders articulated their understanding of forms of colonisation in the EPI. They acknowledged that, while these practices were often subtle, they centralised power in the hands of GN funding organisations, thus limiting local agency. This observation echoes historical literature from colonial Cameroon, which depicts a scenario where the indigenous population lacked the free will to consent to clinical trials and vaccinations during health campaigns [4].

### Positive Expressions of Dependence on Global North Funding and Technical Support

Interestingly, some stakeholders viewed the role of GN organisations in a more positive light, highlighting their contributions in providing funding and technical support that have helped reduce morbidity and mortality related to VPDs, particularly during the COVID-19 pandemic. Notably, achievements in GH, such as the eradication of smallpox, are often attributed to the significant efforts of GN organisations within the GS. However, this positive perspective comes with caveats. It may foster a dependency attitude where GS populations perceive themselves as vulnerable and view white individuals as saviours coming to rescue them from their challenges. Such a viewpoint risks reinforcing a colonial mindset of dependency, thereby perpetuating power imbalances and undermining self-determination [40].

#### Institutional Preferential Treatment and the Expatriate Paradox

Several stakeholders, particularly EPI managers and civil society actors, perceived that GN actors involved in Cameroon’s EPI received preferential treatment compared with locally employed staff, notably higher pay for comparable roles. Some participants extended this observation, suggesting that such disparities reflect underlying financial incentives embedded in the structure of development programming itself, rather than incidental or neutral differences in compensation. This perception aligns with the argument that salary and resourcing disparities in development programmes can reflect structural financial incentives favouring GN-affiliated actors and institutions over local counterparts [41]. Similarly, a major interdisciplinary study led by Massey University and Trinity College Dublin found that expatriate aid workers earned, on average, four times more than local employees for comparable work—a gap not explained by differences in experience or skills [42]. Dual salary systems that perpetuate dominance and injustice undermine local workers’ pride and ultimately produce poverty effects rather than capacity-building in the GS [42].

### Challenges and opportunities in relation to decolonising Cameroon’s EPI

#### Dependence on GN funding

In Cameroon, funding for the EPI is predominantly provided by GN organisations, while the government plays a minimal role, a trend common in GN-GS collaborations [43]. Stakeholders indicated that such funding systems perpetuate colonisation by giving GN countries the advantage of influencing decisions to suit GN interests over the priorities and preferences of the local population. One stakeholder described how funders dictate the timelines and strategies for implementing catch-up vaccination interventions. This often leads to suboptimal results in the local context, causing frustration among national actors. Similarly, during the COVID-19 pandemic, the rollout of GN-funded prevention and control measures, such as vaccines, rather than the use of traditional remedies, met with stiff resistance and distrust in Cameroon [15]. Furthermore, stakeholders criticised the Cameroonian government for complacency regarding colonial influences, as evidenced by its minimal financial investment in healthcare. Cameroon allocates less than 5% of its state income to healthcare, significantly below the 15% benchmark established by the Abuja Declaration [44]. Notably, partners like GAVI provide over 85% of the funding required for the EPI. Fortunately, Cameroon is preparing to transition away from GAVI support [18], which could mark a crucial step towards greater independence from GN aid. The recent funding cuts from the USA to the GS present an opportunity to reduce reliance on Western financial support and instead focus on local resource mobilisation strategies to maintain immunisation efforts.

#### The COVID-19 pandemic and Epistemic Injustice

Stakeholders identified the COVID-19 pandemic as a prime example of modern colonisation, consistent with existing literature. The term “vaccine apartheid,” coined by the WHO Director-General, illustrates the widening gap in access to COVID-19 vaccines between Global North (GN) and Global South (GS) countries, which hindered global vaccination efforts [45]. By February 2022, Europe had discarded 55 million COVID-19 vaccine doses—25 million more than the 30 million doses donated to Africa during the same period. Furthermore, the European Union rejected proposals allowing Africa to produce its COVID-19 vaccines [41]. During the COVID-19 pandemic, the global health response featured Western biomedical solutions, particularly vaccines and pharmaceutical interventions. This dominant narrative often led to the marginalisation and even active discrediting of traditional medicine practices from the GS, such as Madagascar’s Covid-Organics [15]. Moreover, despite their long history of use and cultural significance, African these treatments were frequently dismissed as unscientific or ineffective by international health organisations during the pandemic [16]. Meanwhile, resources and funding were channelled towards several vaccine candidates, promoting significant gains for GN-based pharmaceutical companies [46]. Nonetheless, traditional remedies remained a local preference in Cameroonian societies during the pandemic while the roll-out of COVID-19 vaccines faced high rates of vaccine hesitancy [15]. Thus, the interplay between global health responses and the marginalisation of indigenous practices highlights the complexities of the pandemic and its broader implications for health equity.

#### Local vaccine production capacity

Stakeholders indicated the absence of local vaccine manufacturing companies in Cameroon and most African nations as an enabler for continuous dependence on foreign vaccines and a significant barrier to achieving health sovereignty. While VPDs are prominent in the GS, most vaccine manufacturing companies reside in the GN. In Africa, five countries are involved in vaccine manufacturing: Egypt, Morocco, Senegal, South Africa, and Tunisia, with production capacity that does not suffice to cover the needs of the African population and vaccine costs that are too high for low-resource countries [47]. As initially indicated, donating and using Western-developed vaccines in the GS with little or no quality assurance checks are prime factors contributing to vaccine hesitancy. African countries have prioritised enhancing their local vaccine manufacturing capacity in response to the vaccine disparities revealed during the COVID-19 pandemic.

Collaborations such as the African Vaccine Manufacturing Initiative (AVMI) and the African Union’s Partnership for African Vaccine Manufacturing have spurred initiatives to produce vaccines domestically. As a result, countries like Senegal, Rwanda, and South Africa have established vaccine production facilities. This development reduces dependence on Western nations and enhances regional self-sufficiency [47]. Furthermore, it signifies a crucial step toward decolonising health in Africa, as it ensures that the continent can secure access to essential vaccines without being constrained by external supply limitations.

#### Colonial Curriculum and Epistemic Injustice

One stakeholder critiqued the use of an outdated “colonial curriculum” in health training, introduced during the colonial era, requiring epistemic decolonisation. Where educational frameworks continue to prioritise Western medical knowledge above all other forms, they fail to prepare healthcare professionals for their local contexts and discourage the mainstreaming of local knowledge [48]. This sustains a form of knowledge colonialism, where dominant Western epistemologies marginalise or erase alternative ways of knowing, healing and communication that are not grounded in cultural norms. In the related literature, calls to decolonise GH have also focused on decolonising epistemologies so that Indigenous peoples’ voices are heard and traditional practices valued to the same extent as biomedical knowledge. For example, universities in Bolivia and Ecuador have adopted curricula based on Indigenous knowledge and languages, challenging traditional Eurocentric educational models. In Bolivia, the introduced concept of Vivir Bien (‘Living Well’) is based on Indigenous philosophies and connected to environmental and community health. This idea is now integrated into the curriculum, aligning educational objectives with Indigenous cultural values [49]. Similarly, culturally sensitive education will bridge the gap between biomedical and traditional medicine, demonstrating the relevance of both. These changes reflect a move toward a more inclusive, culturally relevant education system honouring Indigenous identities and knowledge.

#### Decentralised governance structure

Stakeholders perceived that the EPI operates through centralised governance systems, in which decisions are made at the central level and then rolled out for implementation at the periphery, without consideration for the communities that benefit from the interventions. This structure of centralised decision-making is known to have been established by colonial powers (French) and inherited by governments following the country’s independence [9]. By contrast, stakeholders cited decentralised governance as a means of empowering local authorities and communities, thereby challenging a key feature of colonial governance [9]. Despite its under-implementation, Cameroon has a decentralisation policy intended to transfer authority and resources from the central government to regional and local levels. This policy promotes greater citizen participation and improved access to essential services. Established in the 1996 Constitution and further detailed through laws in 2004 and 2008, it seeks to devolve responsibilities across economic, social, health, educational, cultural, and sports development sectors [50]. Adopting decentralised policies will enhance the involvement of local non-health stakeholders in health interventions, including the financing and sustainability of immunisation efforts. Decentralisation is also necessary for designing and implementing community-tailored immunisation approaches that reflect the needs and priorities of communities. This philosophy is crucial in Cameroon’s multicultural and bilingual environment, where communities’ priorities and needs vary within and across regions. Through the implementation of culturally relevant approaches, communities are more likely to accept immunisation, reducing vaccine hesitancy rates and enhancing EPI vaccination uptake. However, implementing decentralisation does not automatically imply change, nor does it mean that local entities can effectively manage the system for which they are responsible. As an Indonesian study indicates, decentralisation policies must be deployed with capacity-building initiatives that improve skills in planning, budgeting, and utilisation to avoid worsening inequities instead of addressing them [51].

#### Participatory approaches

Participatory approaches, such as Human-Centred Design (HCD), enable communities to engage in planning and designing immunisation sessions to meet community needs. This approach harnesses creativity and empathy while promoting active participation and collaboration among stakeholders at all levels. The research uses persona models featuring fictional characters to represent key participants’ immunisation needs, values, aspirations, capabilities, and constraints [52]. Piloted by a local CSO in two health districts in the Southwest region of Cameroon, HCD has strengthened collaboration between health workers and the community and increased vaccine demand in those districts [52]. Results from this pilot can inform policy reform, and the approach can be scaled nationwide to improve community participation in immunisation sessions.

#### Local resource mobilisation

Although the contributions of international organisations to immunisation efforts in the GS are widely acknowledged in the literature, there is comparatively limited attention to domestically mobilised resources, beyond the well-documented role of community members in social mobilisation. To our knowledge, local philanthropists contribute to immunisation ventures by donating resources, like funds, vehicles, and personal protective equipment, to immunisation actors, yet journals rarely capture these. Moreover, predominance in GN funding may shape what gets studied and published as much as what actually happens on the ground [53]. Reducing reliance on external support requires mobilising resources at all levels of Cameroon’s national system to advance immunisation initiatives sustainably. This aligns with a wider continental push for African governments to increase health financing and attain sovereignty, particularly as nations transition from GAVI support to their immunisation programmes [54]. Our findings also suggest reinvesting funds generated by health facilities into immunisation interventions in government-owned budget lines rather than donor-dependent allocations.

#### Streamlining Local Knowledge

There is increasing acknowledgement of the advantages of integrating local health knowledge, community health workers, and traditional healing methods into health systems collaboratively [55,56] Valuing and recognising local expertise can boost community involvement, make immunisation programmes more culturally suitable, and create connections between biomedical and traditional healthcare systems. Stakeholders suggest that working collaboratively with traditional birth attendants, who are key gatekeepers in many rural communities, will grant access to many vaccine-hesitant communities and increase vaccine uptake. Regulating the traditional medicine sector and effectively communicating the advantages of both traditional medicine and vaccination can enhance the representation of traditional practices. In Mali, traditional medicine is structured under a dedicated department that facilitates regular training and knowledge-sharing sessions between unconventional and traditional health practitioners. This collaborative approach recognises the important role of traditional medicine in achieving UHC goals [55].

### Limitations

My findings are based on the perspectives of stakeholders in Cameroon’s Southwest region and may not represent the views of EPI stakeholders nationwide. A significant limitation is the overrepresentation of health professionals, who compose 73% of the sample. This may have skewed the primary themes, resulting in a narrow perspective. Future research should aim for a more balanced representation of participants, including beneficiaries and policymakers, to enhance validity.

### Conclusion

Our study contributes to the limited body of knowledge on decolonial studies in Africa, offering novel insight into stakeholders’ views on Cameroon’s EPI. The analysis reveals that participants hold complex, sometimes contradictory views on colonisation within the EPI. While acknowledging the advantages of GN funding and technical support, they also recognise the structural inequities that such relationships can perpetuate. These include insufficient government investment in health, centralised governance structures, disparities in compensation between expatriate and local staff, and the marginalisation of indigenous knowledge systems. Moreover, our findings articulate proposed approaches towards a more decolonised EPI, including mobilising local resources, adopting decentralised and participatory programme design, investing in local vaccine manufacturing capabilities, and integrating traditional health knowledge alongside biomedical practices. Consistent with the study’s exploratory design, the findings are diagnostic and map stakeholder perspectives that can inform policies promoting equity, local ownership, and programme sustainability. This research is especially timely as Cameroon transitions away from GAVI support and advances its decentralisation and UHC agendas, moving from an externally funded, centrally governed structure towards a more nationally financed and locally governed health system. As Fanon reminds us [57], however, decolonisation is a complex and continuous process; patience and persistence are required, as immediate transformation is unlikely to materialise. Given the Southwest region’s distinct Anglophone colonial and post-colonial trajectory, future decolonial research in the EPI should target Cameroon’s Francophone regions to determine whether the perceptions documented here reflect a broader national pattern or region-specific historical and political dynamics. Further work should also incorporate the perspectives of immunisation beneficiaries and their caregivers, whose experience of the EPI as recipients may surface further insights not captured by the stakeholder groups examined here.

## Data Availability

The authors report no legal or ethical concerns regarding this study.

## Acknowledgments

To all immunisation actors in Cameroon, braving the odds to ensure no child is left behind.

## APPENDICES

## Appendix 1: Semi-Structured Interview Guide

## Introduction

Welcome, and thank you for participating in this interview. This interview aims to explore stakeholders’ perceptions, opportunities, and challenges within the Expanded Programme for Immunisation (EPI) in the Southwest region of Cameroon through a decolonial lens. Your insights and experiences can help shed light on the role of colonial legacies in health programmes in Cameroon and may contribute to developing more equitable and locally grounded approaches within the EPI.

This interview will be conducted respectfully and will ensure anonymity whereby all identifiers like participant name, place of work, gender and any information that can easily lead to the identification of the participant will be removed and unique identifiers will be used. Confidentiality will be maintained through the secured storage of interview transcripts in OneDrive, only the research team will have access to this information.

Also, consent forms will be stored separately from transcripts to prevent participant identification. Participation is voluntary; you can refuse to answer questions or withdraw from the study anytime. I appreciate your willingness to engage in this discussion; your contributions will be insightful for this study.

### I. Background Information

I-a. At what level do you work in (with) the EPI?

I-b. How long have you worked in (with) the EPI?

I-c. What is your highest level of education completed?

### II. Understanding Coloniality and Decolonisation

Let me tell you a short story to give some context about what I mean by ‘coloniality’ in global health, as this will be a core topic that we will explore together in this interview:

*The Cameroon-Belgium Cooperation in Health is a prime example of coloniality in global health. Established in the 1950s, this cooperation program aimed to improve healthcare infrastructure and services in Cameroon, but Belgian interests and priorities drove it. One of the program’s key components was establishing a network of health facilities and hospitals built and managed by Belgian experts. While these facilities provided much-needed healthcare services, they were also seen as a symbol of Belgian colonial power and influence. In addition, the program relied heavily on Belgian medical personnel, who were often more qualified and better paid than their Cameroonian counterparts. This created a power imbalance, where Belgian experts significantly influenced healthcare decision-making in Cameroon*.

II-a. How does this story relate to, or not relate to, how you think about the colonial influences that are still in place today in Cameroon?

- Can you share a personal experience that shaped your understanding of this?

II-b. What does the idea of decolonisation mean to you, to global health practices in Cameroon?

- In what ways do you think the colonial history of Cameroon is shaping current global health practices (if at all)

- Are there any specific practices or policies related to global health that you think need re-evaluation?

II-c. In what ways are Cameroon’s EPI still shaped by colonial power dynamics, if at all?

- Probe related to policy, funding, decision-making, vaccine delivery, and implementation.

### III. Experiences and Challenges

III-a. What are your most significant challenges in implementing EPI policies?

- Can you provide any examples, based on your personal experience?

- Probe related to: policy, implementation, logistics or decision-making.

- Probe related to recent funding cuts

III-b. How do these challenges relate, or not relate, to the colonial history and power dynamics in the country?

- Probe related to any colonial dynamics associated with recent funding cuts

III-c. Do you see any opportunities to work toward changing these dynamics in these practices?

- Probe related to policy, funding, decision-making, vaccine delivery, and implementation.

- Can you give examples of any current practices in your region that potentially play a role in dismantling colonial legacies?

### IV: Power Dynamics and Partnerships

IV-a. Who are the primary stakeholders in the EPI decision-making process?

- Which stakeholders are often left out, and is this shaped by colonial dynamics or unrelated?

Probe related to: programme beneficiaries, CSOs

- How do the interests and perspectives of those included in decision-making shape EPI’s priorities?

IV-b. How can partnerships between international organisations and local stakeholders be improved?

- What practices could promote mutual respect and accountability in shared decision-making?

- In what ways might the programme change if there were a more balanced representation of voices at the decision-making table?

### V. Opportunities for Decolonisation

V-a. What steps could be taken to work toward local ownership of the EPI?

- Probe related to: advocacy, policy, funding, implementation or decision-making

V-b. How do you think local ownership could enhance the effectiveness of the EPI?

- How could local knowledge and traditional practices be integrated into the EPI?

- What changes do you envision in health outcomes, if any?

### VI. Conclusions & Wrapping Up

VI-a. Are there any other insights you can share about how the EPI could work toward being ‘decolonised’, and what this might look like if completed?

Thank you for participating in this interview. Your time and input are invaluable to this research.

## Appendix 2: Study Participant Information Sheet

Study Title: Drawing on a Decolonial Lens to Explore Stakeholder Perceptions, Opportunities, and Challenges in the Expanded Programme for Immunisation in Southwest Region, Cameroon: A Critical Qualitative Case Study Analysis (V1.0 13.05.2025) The purpose and value of the study. Thank you for considering participation in this research study. This study aims to use a Decolonial lens to explore stakeholders’ perceptions, opportunities, and challenges in the EPI in Cameroon. Your participation will provide valuable insights into the experiences and perspectives of stakeholders involved in the EPI.

Oben Pamela Ayuk-Maru epse Besong, a Master’s in Global Health Student at the Liverpool School of Tropical Medicine (LSTM), U.K., organises the study.

You are invited to participate in a study on “Drawing on a Decolonial Lens to Explore Stakeholder Perceptions, Opportunities, and Challenges in the Expanded Programme for Immunisation in Southwest Region, Cameroon: A Critical Qualitative Case Study Analysis.” The interview will explore your perceptions and knowledge of opportunities and challenges within the EPI, which is critical in closing the knowledge gap on Decolonising Global Health in Cameroon.

You have been considered for this study due to your position as a valued and experienced stakeholder who can provide insights on the topic researched. Your insights and experiences can help shed light on the role of colonial legacies in health programmes in Cameroon and may contribute to developing more equitable and locally grounded approaches within the EPI.

After being informed about the survey, you will have ten days to confirm your participation.

If you agree to participate in the study, you will participate in a 45-90-minute online interview on the study topic. The researcher will use a semi-structured interview guide. You will determine the time and date of the interview, and it will be audio- and video-recorded.

You may experience emotional distress when sharing your experiences and recounting painful memories during the interview. To address this, Participants will be informed of the potential risk due to participation, and only those who consent will be included in the study. The PI will also show empathy and understanding and take breaks when distress occurs.

Your participation in this study is entirely voluntary, and there are no consequences for refusing to participate.

You can withdraw anytime after granting consent to participate in the study. You can cancel your participation by emailing, calling, or verbally informing the PI. The study will not include all documentation of withdrawn participants and will be deleted or burned. Withdrawals are possible up to two weeks following the participants’ interview.

To ensure privacy and anonymity:

All participant information will be collected using unique identifiers rather than personal identification to maintain anonymity. The PI will ensure participants’ names, posts of responsibilities, and job locations are not included. Consent forms will be secured separately from data responses, ensuring that participants cannot be traced back to their responses.

During the analysis phase, data will be aggregated and de-identified to prevent the identification of individual participants.

Study participation does not involve remuneration or financial benefit. However, it will contribute to a better understanding of coloniality and decolonisation within the EPI, Cameroon, and inform strategies for developing a decolonising framework in the EPI.

Safeguarding:

All collected data will be anonymised and stored in OneDrive. Only the PI will access the raw data, and data analysis will be conducted within secure and restricted environments. All collected data will be stored in encrypted digital files accessible only to the research team.

Regular backups will be implemented on OneDrive to prevent data loss while ensuring security.

The collected data will be disseminated as follows;

- Used to elaborate a Master’s dissertation for the obtention of an MSc in Global Health at LSTM, U.K.
- Presented at local scientific conferences in Cameroon and the LSTM Decolonising Global Health conferences, offering a valuable platform for extensive academic dialogue and engagement with experts in the field.
- Published in peer-reviewed journals to ensure accessibility and scrutiny by the academic community. To enhance visibility and reach, consideration will be given to open-access publication options.
- After the research, the findings will be shared with study participants, giving them a meaningful interpretation of their data. Additionally, the results will be communicated to the Ministry of Public Health through the EPI to inform potential policy changes.

Contact for further information

- Oben Pamela Ayuk-Maru epse Besong Phone: (+237) 674941390
- Master’s Research Panel (MRP) contact details (

## Appendix 3: Consent Form

Principal Investigator: Oben Pamela Ayuk-Maru, epse Besong

Study Site: Southwest region, Cameroon

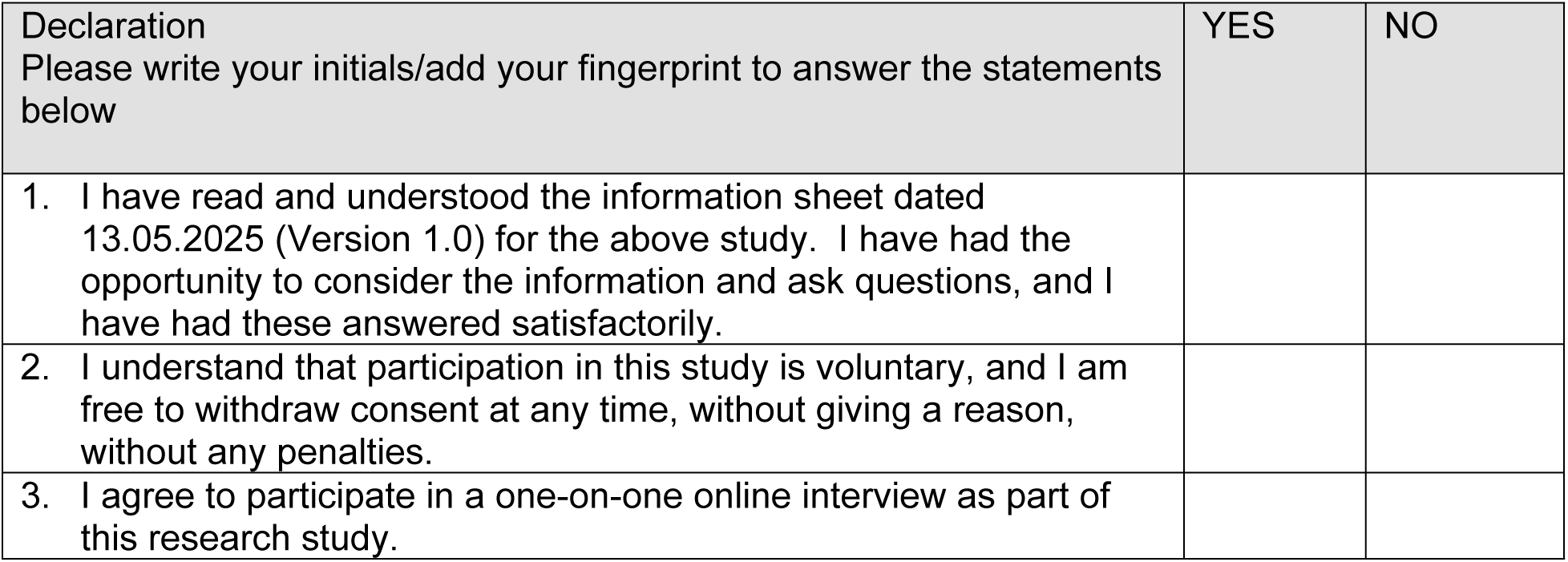
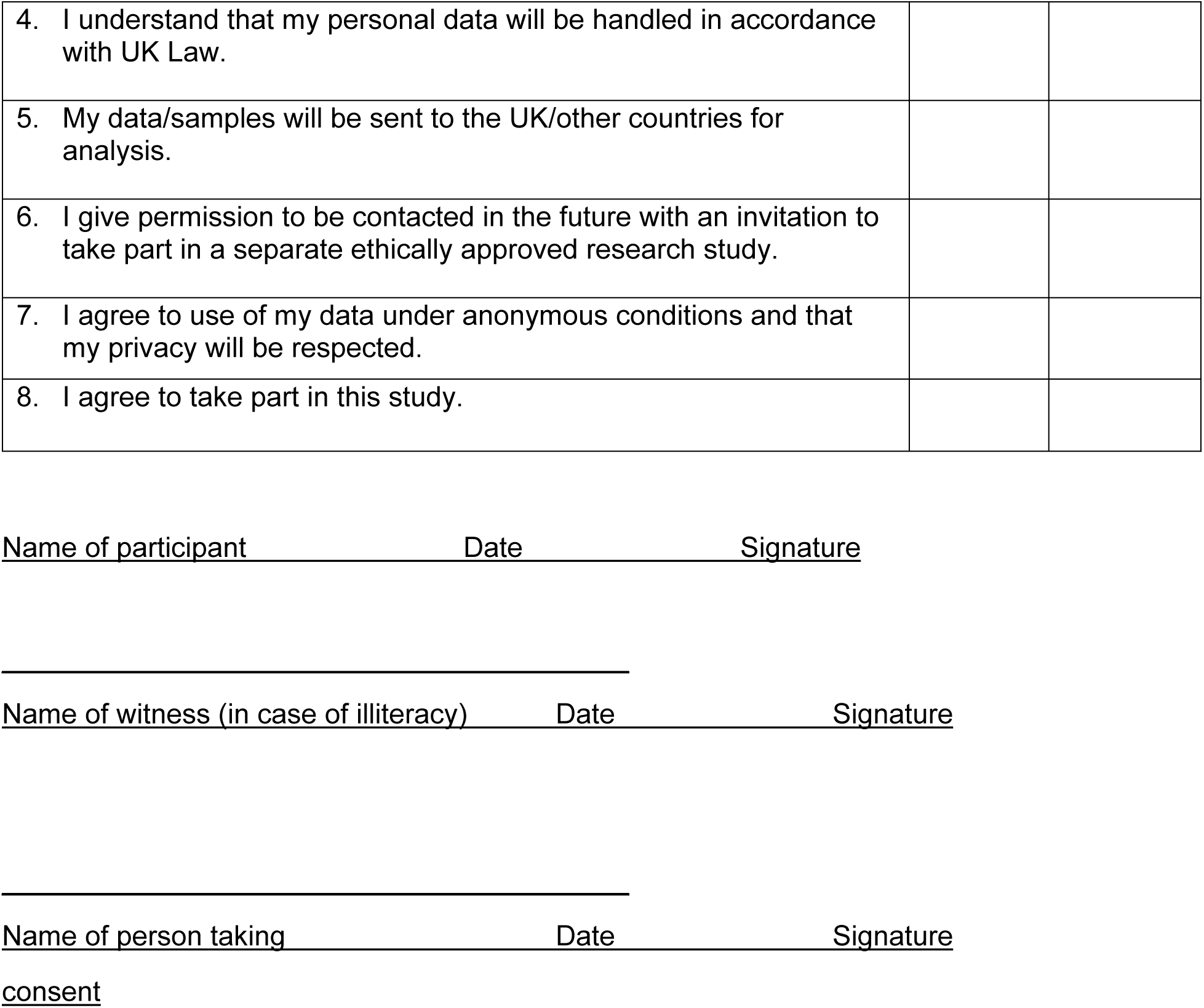

## Appendix 4: Southwest Regional Ethics Committee for Human Health Research in Cameroon Ethical Clearance

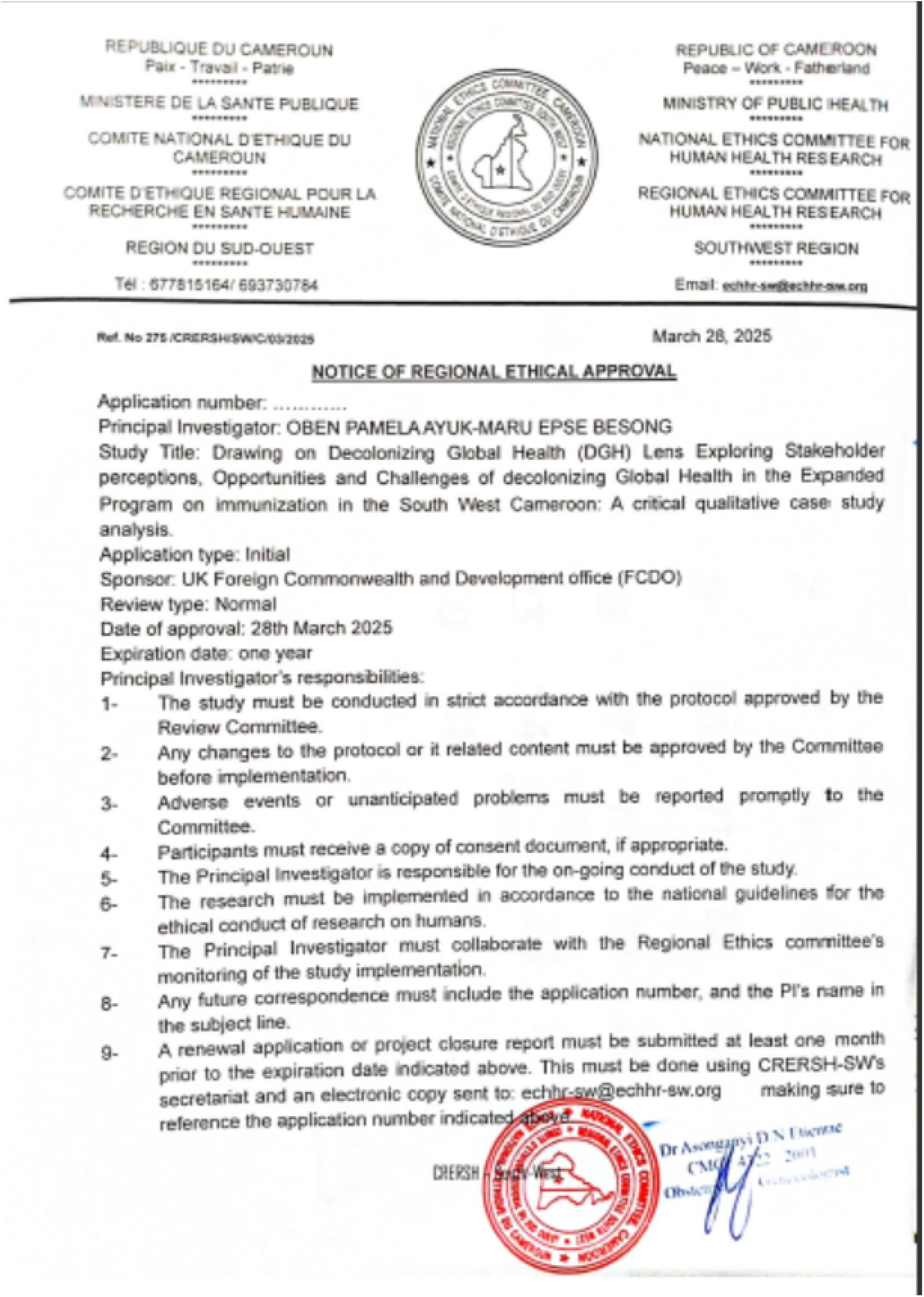

## Appendix 5: LSTM Master Review Panel Ethical Clearance

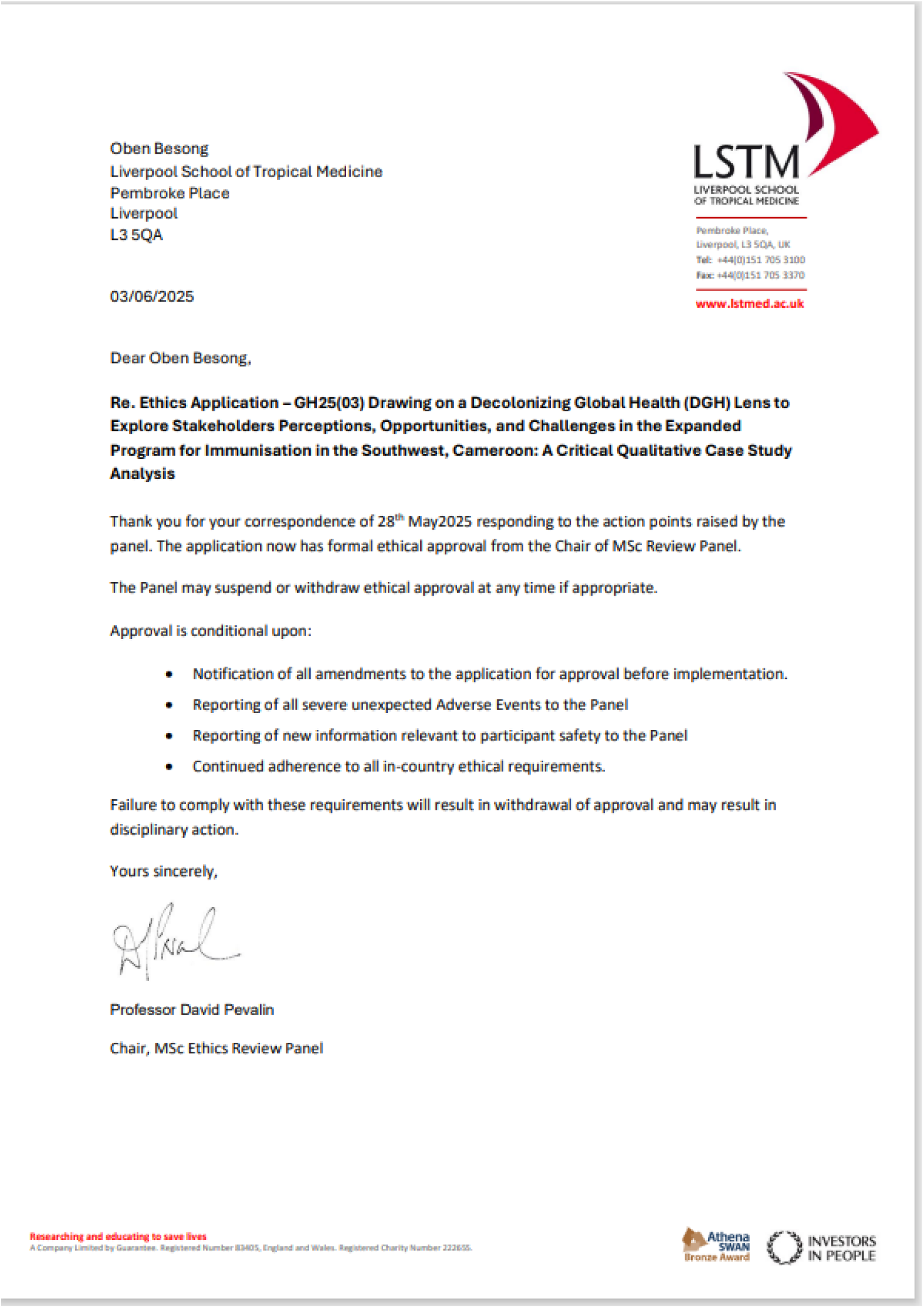

## Appendix 6: NVIVO codes

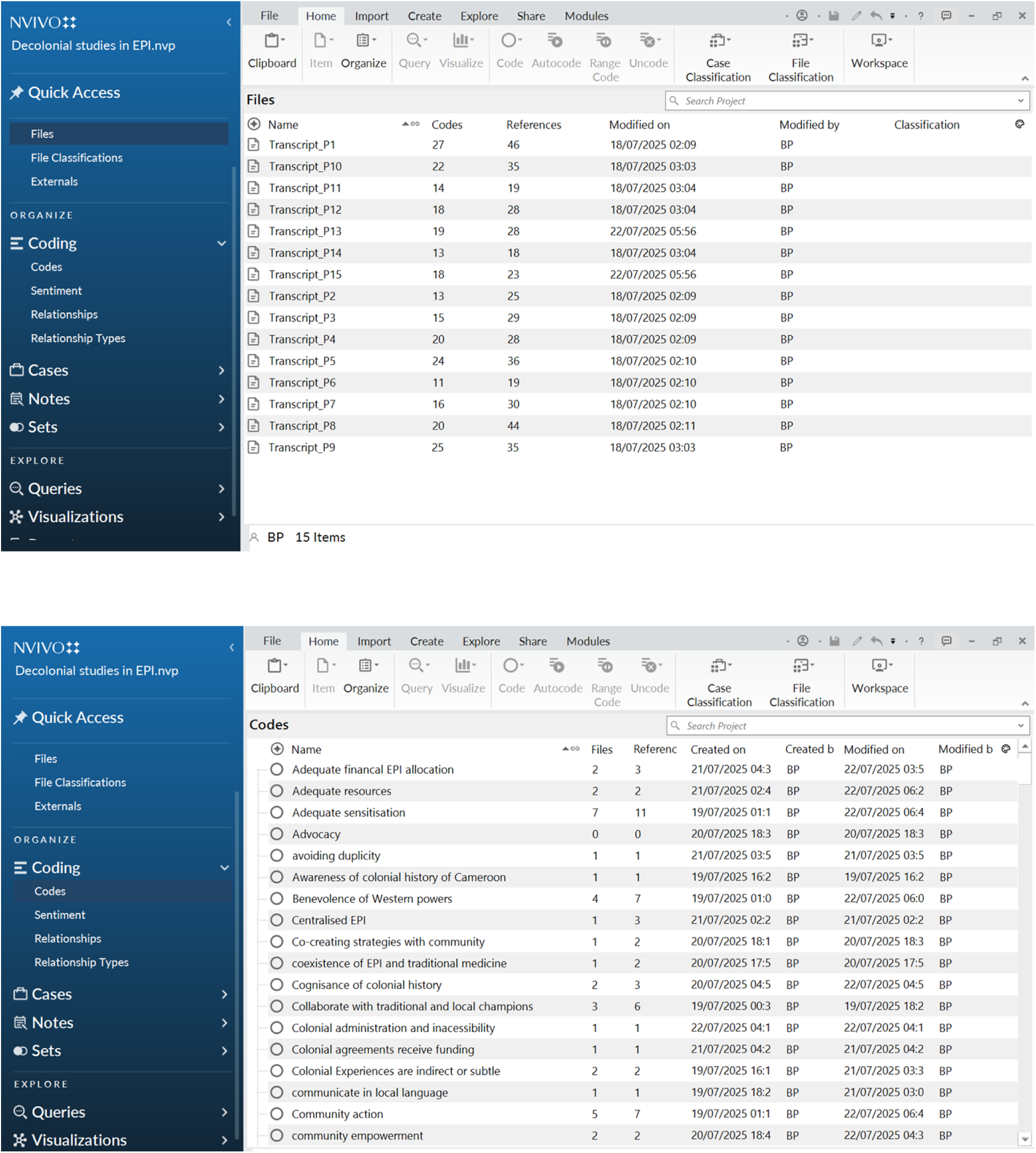

